# Cultural and linguistic adaptation of a personalised post-diagnostic dementia intervention for Bangladeshi families in the UK: an iterative adaptation study

**DOI:** 10.64898/2026.10.13.26365344

**Authors:** Jessica Budgett, Iain Lang, Anwesha Sen, Maisha Hussain, Melisa Yilmaz, Margaret Ogden, Rosemary Philips, Sara Banks, Victoria Vickerstaff, Jana Mikova, Yvonne Birks, Claudia Cooper

**Affiliations:** Centre for Psychiatry and Mental Health, Queen Mary University of London, London, UK; Department of Health and Community Sciences, University of Exeter Medical School, Exeter, UK; National Institute for Health and Care Research Applied Research Collaboration: South West (NIHR ARC South West), UK; Alzheimer’s Society and INSTILLED Patient and Public Involvement (PPI) Group, London, UK; Centre for Evaluation and Methods, Wolfson Institute of Population Health, Queen Mary University of London, London, England, UK; East London NHS Foundation Trust, London, UK; School of Business and Society, University of York, UK

## Abstract

People from UK South Asian communities experience barriers to accessing dementia care. Evidence suggests that evidence-based psychosocial dementia interventions can be successfully adapted for different cultural contexts, but further evidence is needed on how adaptations are developed and refined in practice. We aimed to adapt GRACE-family (Goal diRected, Accessible and Evidence-based Care), an evidence-based, manualised post-diagnostic dementia intervention, comprising 4-8 sessions for family carers and where possible, people with dementia, for Bengali-speaking communities.

In a theory-informed process, a co-adaptation group encompassing lived, academic and clinical/professional experience developed consensus regarding which intervention functions were core. They then explored how adaptation might, while retaining these, account for cultural and linguistic contexts, informed by focus groups with memory service and Bengali community organisation staff (n=14). We developed a prototype adaptation, then piloted it with four Bangladeshi families supporting a person with dementia. Post-intervention, participants were asked about goal attainment, intervention acceptability, and qualitatively interviewed. Pilot data and a focus group with Sylheti-speaking family carers informed further refinements (n=6).

Across data sources, two themes informed adaptation: (1) stigma, trust and community networks shaped access to dementia support; (2) flexible delivery was needed to respond to diverse language, communication and cultural needs. 3/4 pilot families completed the prototype intervention, which was in English as initial consultation cautioned against developing a written manual in a predominantly spoken language. Participants reported improvement on 7/8 personalised goals set, and all considered the intervention acceptable. Pilot delivery identified challenges using English-language materials during interpreter-mediated sessions. Iterative refinements added standard Bengali materials and spoken Sylheti audiovisual resources to broaden delivery options and facilitator training for culturally responsive and interpreter-mediated delivery.

Our co-adaptation approach enabled stakeholders to advise from different, including experiential perspectives. It may inform future approaches to cultural and linguistically adaptations of psychological interventions for minoritised communities.

## Introduction

Dementia is a leading cause of disability and dependency worldwide, with over a million people living with dementia in the UK (Carnell Farrar, 2024). Most people wish to remain living at home for as long as possible (Rapaport et al., 2020), yet many experience fragmented post-diagnostic care, unmet psychosocial needs and limited access to evidence-based support (Prince et al., 2014). National policy increasingly emphasises the need for equitable, personalised, community-based dementia care that enables people to live well with dementia (Department of Health and Social Care, 2025; NICE, 2018).

Despite this, access to evidence-based post-diagnostic support remains inconsistent, particularly for people from minority ethnic communities (Bhanu et al., 2020; Carter et al., 2024; Herat-Gunaratne et al., 2020; Penfold et al., 2026). Dementia prevalence in South Asian communities, the largest minority ethnic population in England, is expected to increase significantly over coming decades (Carter et al., 2024; Mukadam et al., 2022). Barriers South Asian UK communities experience when accessing formal dementia care, include: stigma surrounding dementia, language barriers, limited culturally appropriate services, difficulties navigating care pathways and poor statutory and community organisational integration (Armstrong et al., 2019; McDermott et al., 2025; Nair et al., 2026). These barriers contribute to inequalities in access to diagnosis, post-diagnostic support and care (Cheston et al., 2025).

Evidence-based psychosocial interventions can improve outcomes for people diagnosed with dementia and their family carers, but most have been developed and evaluated in predominantly White populations (Kishita et al., 2018). Cultural and linguistic adaptation aims to improve relevance and accessibility across under-served populations and settings, through changes to intervention content, communication and delivery. Evidence suggests that established psychosocial dementia interventions can be successfully adapted for different cultural contexts while retaining their core therapeutic components (James et al., 2021; McDermott et al., 2025). However, the evidence for culturally adapted dementia interventions remains limited, including for post-diagnostic support for South Asian communities (Akhtar et al., 2026; James et al., 2021). Further evidence and transparent reporting of this adaptation process is needed, including how different perspectives inform decisions, implementation experience informs refinement, and core intervention functions are retained. This may inform adaptation of established interventions to other cultural and linguistic contexts (James et al., 2021; Moore et al., 2021; Wiltsey Stirman et al., 2019).

GRACE-family (Goal Directed, Accessible and Evidence-based Care), formerly known as NIDUS-Family (New Interventions for Independence in Dementia Studies), is a manualised, modular psychosocial post-diagnostic dementia care intervention. Facilitators work with people living with dementia and their family carers to identify personalised goals using Goal Attainment Scaling (GAS), a structured method for defining and evaluating progress towards individually meaningful outcomes. Facilitators then deliver up to eight structured support sessions selected from a menu of modules to align with the dyad’s goals, then two to four brief follow-up support calls over 12 months. The GRACE-family modules and materials can be accessed via the website: https://qmwiseminds.co.uk/grace/. In a multicentre Randomised Controlled Trial (RCT), GRACE-family improved personalised goal attainment and was clinically and cost-effective compared with goal setting alone (Cooper et al., 2024). The intervention was designed to be scalable, with trained non-clinical facilitators (such as social prescribers and dementia advisors) delivering structured, person-centred support using manualised resources.

Although GRACE-family is designed to be personalised to individual and family priorities, its materials, facilitator training and delivery processes were developed and evaluated predominantly within English-speaking UK services. Cultural and linguistic adaptation was therefore needed to support its accessible and meaningful delivery with Bengali-speaking families.

We aimed to adapt GRACE-family to support culturally and linguistically responsive delivery with Bangladeshi families in East London, home to one of the largest

Bangladeshi communities in England (Tower Hamlets Partnership, 2023), using a co-produced, iterative adaptation process. The objectives were to:

1. identify the core and adaptable functions of GRACE-family;
2. identify cultural, linguistic and contextual considerations relevant to adapting GRACE-family for Bangladeshi families; and
3. develop and iteratively refine adaptations to GRACE-family content, delivery and facilitator preparation, informed by stakeholder engagement and pilot delivery, and document how and why these adaptations were made.

## Methods

### Study Design

This mixed methods adaptation study followed the ADAPT guidance (de Boer et al., 2025). The Consolidated Framework for Implementation Research (CFIR) (Reardon et al., 2025) informed exploration of contextual influences relevant to adaptation and delivery, and we used the Framework for Reporting Adaptations and Modifications-Enhanced (FRAME) (Wiltsey Stirman et al., 2019) to document adaptations made throughout the process.

London -Camden & Kings Cross Research Ethics Committee (24/LO/0843) approved the study on 9 December 2024. All participants provided written or audio-recorded informed consent before taking part. Where a person living with dementia lacked capacity to provide informed consent, a personal consultee declaration was completed in accordance with the Mental Capacity Act (England and Wales), 2005. Family carers unable to provide informed consent were excluded.

### Co-adaptation and PPI advisory groups

A multidisciplinary co-adaptation group comprising people with lived, professional and academic expertise met seven times between September 2024 and May 2026 to steer the adaptation process. It was co-chaired by IL (a public health and implementation researcher) and JB (Chief Investigator of INSTILLED). Members included CC (Chief Investigator of the NIDUS-Family trial and old age psychiatrist), YB (Professor of Applied Health and Social Care Research), VV (methodologist), JM (senior memory service nurse), a dementia third-sector organisation representative, and two PPI contributors (MO and RP). The group remit was: defining intervention core components, contributing to the development of focus-group topic guides and recruitment strategies, reviewing emerging findings, and considering and refining adaptations arising from stakeholder engagement and pilot delivery. Where the group decided to translate materials, they also guided decisions around the process and checking of these components with native speakers.

A separate PPI advisory group comprising four family carers met quarterly throughout the study. The group reviewed participant-facing materials and discussed emerging findings and proposed adaptations. It was chaired by JB, who facilitated communication between the PPI advisory group and wider co-adaptation group. Maintaining a separate forum provided additional opportunities for family carers to contribute lived-experience perspectives throughout the adaptation process.

### Stage 1: Defining the core components

The co-adaptation group met in September and December 2024 to develop consensus regarding the core components of GRACE: that is, which intervention functions should be retained and which aspects of intervention content or delivery could be modified. Group discussions were informed by the published NIDUS-Family findings (Cooper et al., 2024) and programme theory (Wyman et al., 2024), intervention manuals, and facilitator training materials.

Discussions considered facilitator characteristics and training, supervision, session structure, eligibility, goal setting, and intervention content and materials. Where members initially held differing views, these were discussed within the group, with plans to use voting where consensus could not be reached. The group also considered how the intervention manuals need to be adapted for routine delivery, for example, in the RCT of NIDUS-family, goal-setting was a research process, but for routine delivery was embedded in the intervention.

### Stage 2: Identifying cultural, linguistic and contextual considerations

#### Participants and recruitment

In March 2025, we conducted two focus groups, to explore cultural, linguistic and contextual considerations relevant to adapting GRACE for the Bengali population of East London: one with multidisciplinary memory service staff involved in post-diagnostic dementia care of this group; and one with staff from a local Bengali community health organisation. We held a further focus group with Sylheti-speaking family carers of people living with dementia in January 2026 to explore language and communication issues in greater depth and inform subsequent refinement.

Memory service staff were recruited through co-adaptation group members working within the participating memory service, who circulated study invitations to eligible colleagues. Community organisation staff were recruited through the participating voluntary-sector organisation. Family carers were recruited through a Bengali PPI contributor, who approached family carers within her local community networks.

#### Focus group procedures

The co-adaptation group selected constructs from the updated CFIR (Reardon et al., 2025) considered relevant to cultural and linguistic adaptation to inform topic guides for staff (Appendix 1) and family carers (Appendix 2). Discussions explored understanding of dementia and experiences of post-diagnostic support, cultural expectations surrounding care, language and communication needs, intervention materials and delivery, and recommendations for improving cultural relevance and accessibility.

The memory service and community organisation focus groups were conducted in-person within these services and facilitated by JB and MZ. Participants received an information pack introducing the GRACE-Family intervention (Appendix 3) one week before the groups to introduce the intervention. The family-carer focus group was conducted at our university office in Sylheti by two bilingual researchers (MH and AS). They researchers introduced GRACE verbally in Sylheti, enabling unfamiliar concepts to be clarified during the discussion rather than relying on written information.

Focus groups lasted 60-120 minutes, with breaks, and were audio-recorded. English-language groups were transcribed verbatim and anonymised. The Sylheti-language recording was translated and transcribed into English for analysis.

### Stage 3: Developing and piloting the adapted GRACE prototype

Findings from Stage 1 and the pre-pilot stakeholder focus groups informed development of the GRACE prototype evaluated in the pilot. Its development was steered by the co-adaptation group, with all adaptations recorded using FRAME.

#### Pilot participants and recruitment

Bangladeshi families were recruited through the participating East London memory service, between April and June 2025. Eligible participants were self-defined Bangladeshi dyads comprising a person living with dementia and a family carer. People living with dementia were required to have a documented diagnosis of dementia of any type or severity and to be living in the community. Family carers were required to have at least weekly face-to-face or telephone contact with the person living with dementia.

#### Intervention delivery

GRACE was delivered by a trained facilitator (MZ), who completed the adapted GRACE facilitator training programme before intervention delivery and received monthly clinical supervision from a clinical psychologist (SB).

GRACE comprised four to eight sessions according to family need and preference. Sessions could be delivered face-to-face or remotely by video call. Collaborative goal setting was integrated into intervention delivery (by the co-adaptation group), and GRACE modules were selected according to each family’s personalised goals.

#### Pilot measures and qualitative interviews

Baseline demographic information was collected from family carers. Pilot study process data collected included details of recruitment, retention, number of sessions completed, mode and language of delivery, and personalised goals. Goal status was recorded at final review. Acceptability was assessed following intervention completion using the four-item Acceptability of Intervention Measure (AIM) (25; Appendix 5).

Following intervention completion and six-month quantitative follow-up, participating people living with dementia and family carers, together with the intervention facilitator, were invited to take part in semi-structured, individual (or dyadic if they preferred) qualitative interviews. Interviews explored experiences of receiving or delivering GRACE, perceived acceptability, barriers encountered during delivery, and recommendations for further adaptation. Topic guides were developed by the research team using the GRACE programme theory as a conceptual framework and reviewed by the PPI advisory and co-adaptation groups (Appendices 4 and 6).

Three English-language interviews (two family carers and the facilitator) were conducted by JB and one Bengali-language interview with family carer by bilingual researchers AS and MH, 8–9 months after baseline. Interviews were conducted in participants’ preferred language, audio-recorded and transcribed, with the Bengali-language interview translated into English for analysis.

### Procedures for developing translated and audiovisual materials

Where the co-adaptation process identified a need for translated participant-facing materials, standard written Bengali versions were developed using an iterative, meaning-based approach. The booklets were translated by a professional translation service and reviewed by Bengali-speaking PPI contributors. Concepts and phrases without straightforward or natural direct equivalents were discussed with PPI contributors to identify wording that conveyed the intended meaning rather than prioritising word-for-word equivalence. Examples and names were adapted where appropriate to improve cultural relevance.

Where spoken Sylheti resources were agreed as an additional format, a Sylheti-speaking PPI contributor and an interpreter whose first language was Sylheti translated relevant text-heavy content into spoken Sylheti. They presented the translated material to camera for professional filming. Bilingual research assistants reviewed the completed recordings for accuracy and formatting, and videos were accessed through QR codes embedded within intervention materials.

### Analysis

Participant characteristics and pilot recruitment, retention, intervention delivery, personalised goals and acceptability scores were summarised descriptively.

Qualitative data were analysed using reflexive thematic analysis (Braun & Clarke, 2019), supported by NVivo 14. For English-language data, JB and MY familiarised themselves with transcripts and undertook initial coding before comparing and discussing their interpretations. JB subsequently organised codes into candidate themes focused on cultural, linguistic and contextual considerations relevant to adaptation.

For Sylheti-and Bengali-language data, bilingual researchers AS and MH reviewed original-language recordings alongside translated transcripts and discussed the interpretation of culturally and linguistically specific concepts. They identified initial codes and observations before JB undertook thematic coding of the translated data. Emerging interpretations across datasets were discussed with MY, AS, MH and IL to consider alternative interpretations and support appropriate representation of culturally and linguistically specific concepts.

Pilot interview findings were considered alongside descriptive information on intervention delivery, language and mode of delivery, personalised goals and goal status to inform subsequent adaptation.

### Reflexivity

JB, CC and IL were closely involved in the original development and evaluation of the NIDUS-Family intervention and brought detailed knowledge of the intervention and its theoretical foundations. In contrast, MY, AS, MH, YB, JM joined the programme during the INSTILLED adaptation study and contributed perspectives independent of the intervention’s original development. These complementary perspectives informed data interpretation. Emerging findings and proposed adaptations were discussed within the co-adaptation and PPI advisory groups to challenge assumptions and incorporate both empirical evidence and lived experience.

### Documenting adaptations using FRAME

Adaptations agreed and implemented throughout the study were documented using FRAME (Wiltsey Stirman et al., 2019). Documentation captured which evidence or stage of the adaptation process informed each decision, what was adapted and why, whether adaptations were planned before implementation or made reactively in response to implementation experience, and their anticipated implications for intervention fidelity.

Consideration of fidelity was distinct from assessment against the agreed core components. Core components provided a reference point for determining whether functions considered essential to GRACE were retained following adaptation. Separately, adaptations were classified as fidelity-consistent where they were not expected to alter the intervention’s underlying functioning or effectiveness, or fidelity-uncertain where their effect on intervention functioning or effectiveness was unknown. Adaptations and their classifications were reviewed iteratively by the co-adaptation group as GRACE was refined.

## Results

### Objective 1: Defining the core components of GRACE

The co-adaptation group reached consensus on eight core components considered important for preserving the functions of GRACE during adaptation (Table 1). Discussions centred on balancing fidelity to the model evaluated in the trial with pragmatism, while ensuring reach and acceptability across a broad range of participants, services and staff.

**Table 1.** Core components of GRACE used to guide adaptation decisions.

| **Core component** | **Rationale** |
| --- | --- |
| **One consistent facilitator delivers the intervention wherever possible.** | Maintains continuity, therapeutic rapport and trust throughout the intervention. |
| **GRACE is delivered by trained facilitators without requiring specialist clinical qualifications.** | Preserves the intervention's scalability and accessibility across routine services. |
| **Facilitators receive regular supervision from an appropriately skilled supervisor.** | Supports fidelity, reflective practice and safe delivery. |
| **A primary carer/supporter is identified** | Maintains the dyadic, family-centred nature of the intervention |
| **GRACE comprises 4–8 sessions with optional catch-up calls.** | Preserves the structured yet flexible delivery model. |
| **Each dyad identifies at least one personalised goal through collaborative goal setting in session 1** | Preserves collaborative goal setting as the intervention's central mechanism of change. |
| **GRACE modules are selected according to the personalised goals identified with each family across sessions, aiming for 3-5 goals.** | Maintains the goal-directed and personalised nature of the intervention by ensuring that intervention content is selected in response to each family's priorities. |
| **All new intervention modules and substantial modifications are reviewed against the agreed core components before implementation.** | Ensures future adaptations remain aligned with the intervention's core functions while improving contextual fit. |

Intervention dose and goal setting generated discussion because these represented departures from the trial model of 6-8 sessions and 3-5 goals set prior to randomisation. The group considered it important to retain collaborative personalised goal setting while allowing greater flexibility in when goals were set. Members discussed how families might initially have one pressing priority, with further goals identified or refined as they became familiar with GRACE. The group therefore agreed a flexible intervention dose of 4-8 sessions and that at least one personalised goal should be identified in the first session.

The group also considered how goal setting could be integrated into routine service delivery. Clinicians and researchers raised concerns about the time and complexity of completing full GAS alongside facilitators’ routine roles. An abbreviated GAS procedure, informed by previous work on simplified GAS approaches (Turner-Stokes, 2009), was therefore adopted and adapted for the dementia context, with only the baseline (0) and expected (+1) outcome levels initially defined.

Other core components largely retained functions of the trial intervention while allowing flexibility in how these were operationalised in routine services. These included continuity of facilitator wherever possible; delivery by trained facilitators without requiring specialist clinical qualifications; regular supervision by an appropriately skilled supervisor; involvement of a primary family carer or supporter; selection of intervention modules according to personalised goals; and review of new modules or substantial modifications against the agreed core components.

In the trial, supervision was provided by a clinical psychologist or psychiatrist. The group agreed that defining the competencies required for supervision, rather than specifying professional roles, would support delivery across diverse service settings. This could enable appropriately skilled senior care managers, social workers or others routinely supervising non-clinical staff to undertake the role, supported through a ‘train-the-trainer’ model led by the core research team.

### Objective 2: Cultural, linguistic and contextual considerations for adapting GRACE

Three focus groups involving 20 participants were conducted: seven memory service staff, seven community organisation staff, and six family carers. The community organisation group included six Bengali community facilitators, all of whom were themselves members of the Bengali community, and one programme manager. The family carers were all South Asian women and predominantly daughters-in-law of the person with dementia (5/6). Participant characteristics, including age, gender, ethnicity, language, education, employment and accommodation, are presented in Table 2.

**Table 2:** Characteristics of focus group participants.

| *Results are n\* (%) unless specified otherwise* | **Memory service staff (n=7)** | **Community organisation staff (n=7)** | **Family carers (n=6)** |
| --- | --- | --- | --- |
| **Age (years), mean (SD<sup>†</sup>)** | 41 (8.20) | 45.2 (9.46) | 40.5 (7.7) |
| **Ethnicity** |  |  |  |
| White | 4 (57.1) | 1 (14.2) | 0 (0) |
| South Asian | 1 (14.2) | 6 (85.7) | 6 (100) |
| Black | 2 (28.5) | 0 (0) | 0 (0) |
| **Gender** |  |  |  |
| Male | 0 (0) | 0 (0) | 0 (0) |
| Female | 6 (85.7) | 7 (100) | 6 (100) |
| Nonbinary | 1 (14.2) | 0 (0) | 0 (0) |
| **Main Language** |  |  |  |
| English | 4 (57.1) | 4 (57.1) | 0 (0) |
| Other | 3 (42.8) | 3 (42.8) | 6 (100) |
| **Highest educational attainment** |  |  |  |
| Higher degree | 1 (14.2) | 0 (0) | 1 (16.6) |
| Degree or Diploma | 3 (42.8) | 2 (28.5) | 0 (0) |
| PG Diploma | 2 (28.5) | 0 (0) | 1 (16.6) |
| A level (or equivalent) | 1 (14.2) | 3 (42.8) | 4 (66.6) |
| O levels/GCSE (or equivalent) | 0 (0) | 2 (28.5) | 0 (0) |
| **Employment** |  |  |  |
| Full – time | 6 (85.7) | 1 (14.2) | 0 (0) |
| Part – time | 0 (0) | 5 (71.4) | 1 (16.6) |
| Not Employed | 0 (0) | 0 (0) | 5 (83.3) |
| Prefer not to disclose | 1 (14.2) | 1 (14.2) | 0 (0) |
| **Accommodation** |  |  |  |
| Private rented | 2 (28.5) | 4 (57.1) | 0 (0) |
| Social rented | 0 (0) | 0 (0) | 6 (100) |
| Owned | 4 (57.1) | 1 (14.2) | 0 (0) |
| Prefer not to disclose | 1 (14.2) | 2 (28.5) | 0 (0) |
| **Carer relationship** |  |  |  |
| Daughter in law |  |  | 5 (83.3) |
| Other (friend) |  |  | 1 (16.6) |
| **Co-Habiting** |  |  |  |
| Dyad* are co-habiting |  |  | 3 (50) |
| Dyad* live apart |  |  | 3 (50) |

\*Dyad refers to a person living with dementia and a family carer pair

Two themes were identified that informed the cultural and linguistic adaptation of GRACE: (1) stigma, trust and community networks shaped access to dementia support; and (2) flexible delivery was needed to respond to diverse language, communication and cultural needs.

#### Theme 1: Stigma, trust and community networks shaped access to dementia support

Participants described how understandings of dementia, stigma, trust and community networks could influence families’ engagement with dementia support. Community facilitators and family carers described dementia as highly stigmatised among some older members of the Bangladeshi community, with symptoms sometimes understood as part of normal ageing or mental illness. Participants also described difficulties conveying the concept of dementia in Bengali, with one family carer commenting that “*there’s no direct word for it [dementia] in Bengali*” [FC6, family carer].

> *“In Bangladesh, they never diagnose it. They just say he’s gone mad… the community just laughs at it.”* [CF1, community facilitator]
>
> *“People just say God gave them the illness.”* [FC3, Family Carer]

Memory service staff similarly described misconceptions about formal dementia support, including concerns that accessing social care could be associated with loss of independence or institutional care:

> *“They’re often quite surprised to hear about that kind of support and just associate the social work support with going into a nursing home and having your kids not love you anymore.”* [S4, psychiatrist]

All three participant groups highlighted the importance of building trust, increasing awareness of dementia and available support, and engaging families through established community networks. Concerns regarding confidentiality were also raised, particularly where interpreters or support workers came from within the local Bengali community.

> *“…they prefer family because they’re worried about confidentiality… translators come from that very tight-knit community.”* [S7, psychiatrist]

Family carers recommended engaging families through trusted community networks, including community organisations, GP surgeries, mosques and community champions, alongside conventional healthcare referral pathways

> *“Word of mouth is the strongest one, so you need to spread the word… when it comes from a reliable source everyone trusts it.”* [FC2, Family Carer]

Family carers also suggested that engagement strategies should be tailored to different groups within the community. For example, three family carers agreed that involving respected male community members or faith leaders may improve awareness and engagement among men.

> *“The community needs knowledge on dementia… men need to be taught… if they go to the mosque and another man is telling them, hopefully they will listen.”* [FC7, Family Carer]

#### Theme 2: Flexible delivery was needed to respond to diverse language, communication and cultural needs

Participants described considerable variation within the Bangladeshi community in language, literacy, communication preferences and culturally meaningful ways of engaging with support. Rather than identifying a single approach that would meet the needs of all families, their accounts emphasised the need for flexibility in intervention materials and delivery.

During pre-pilot consultation, community facilitators questioned whether translating GRACE materials into formal written Bengali would necessarily improve accessibility. They described many older adults in the local Bangladeshi community as primarily communicating in spoken Sylheti and highlighted differences between spoken and written language:

> *“There is no point in just translating the booklets formally… the formal Bangla script would be like me handing you an English leaflet written in Shakespearean English. Sylheti dialect is just spoken.”* [CF3, community facilitator]

Community facilitators also emphasised that accessibility depended on conveying the meaning of concepts rather than translating terminology literally:

> *“It’s not just translating into any version of Bengali, it’s the words like dementia, intervention… someone has to understand what those mean and explain them in a way people can understand.”* [CF5, community facilitator]

Informed by these concerns, English-language participant materials were retained in the prototype taken forward into pilot delivery, rather than developing a written Bengali translation at this stage.

Subsequent consultation with Sylheti-speaking family carers provided a more nuanced picture of language and communication preferences. Some family carers preferred reading English despite speaking Bengali fluently at home:

“I’m better at reading English than Bengali… I can speak Bengali fluently, but I cannot read.” [FC1, family carer]

However, family carers also highlighted the value of providing information in different languages and formats according to individual preference. Spoken explanations, audio resources and short videos were suggested as potentially more accessible than written information for some families:

> *“Sometimes even Sylheti when you write it down it’s different… speaking is OK… maybe a brief video about the modules.”* [FC1, family carer]

Interpreter-supported delivery introduced additional considerations. Family carers described the importance of matching interpreters to families’ spoken language and dialect, alongside concerns about the quality and consistency of interpretation:

> *“Not everyone is comfortable with an interpreter because sometimes they don’t explain everything properly. Sometimes they send the wrong interpreter as well; sometimes you speak Sylheti, and they send someone who speaks Dhaka Bengali.”* [FC4, family carer]

Memory service staff similarly suggested that interpreters needed to understand

GRACE’s collaborative, goal-directed approach rather than translating individual words or phrases without the wider intervention context. Continuity of interpreter across sessions was also considered important:

> *“It might be good… to have additional training for interpreters around the language that you’re going to be using with your goal setting.”* [S3, Psychologist]
>
> *“It would be good if you could request a consistent interpreter to do all of the sessions instead of having to explain to a new person each session.”* [S3, Psychologist]

Participants also emphasised that culturally responsive delivery extended beyond language. Culturally representative images and examples were considered important for helping families recognise the relevance of the intervention:

> *“If some of the pictures in the booklet wear hijab… we can recognise that this is for the Bangladeshi community.”* [FC2, family carer]

Flexibility in how the intervention was delivered was also considered important for accommodating caring responsibilities, family commitments and practical barriers to attending appointments. Home-based delivery was considered particularly helpful for some families:

> *“[Home visits are] just easier… if you’ve got other kids around and when the person can come to you and work around your little ones, even if you get 30 minutes, it’s much more beneficial than not having it at all.”* [CF6, community facilitator]

Rather than identifying a single preferred model of culturally adapted delivery, participants described differing cultural, linguistic and practical needs and preferences between families. Community facilitators considered GRACE’s personalised, goal-directed structure to provide a basis for responding flexibly to these differences:

> *“The thing is everyone will want a different delivery approach, won’t they?… but its good GRACE fits well into this… the individual[ised] goals help, I think.”* [CF2, community facilitator]

Overall, participants’ accounts indicated a need for a flexible approach in which language, communication format and intervention delivery could respond to individual and family preferences, rather than relying on a single culturally or linguistically adapted version of GRACE

### Objective 3: Developing and iteratively refining adaptations to GRACE

#### Development of the adapted GRACE prototype

Planned adaptations arising from the core-component work were incorporated into the prototype to support delivery in routine services (Table 4).

Pre-pilot focus-group findings also informed cultural adaptation of the prototype. In response to participants’ emphasis on culturally meaningful materials (Theme 2), photographs and illustrations were revised to better represent Bangladeshi families and culturally relevant everyday activities.

The co-adaptation group also considered whether participant-facing materials should be translated before pilot delivery. However, pre-pilot community facilitators strongly cautioned against assuming that formal written Bengali would be more accessible than English, particularly for older people who primarily communicated in spoken Sylheti (Theme 2). The group therefore agreed to retain English-language participant materials for the initial prototype, with interpreter-supported delivery available where required, and to review language and communication needs through pilot delivery.

#### Pilot study findings

The adapted prototype was piloted in 2025. Five eligible Bangladeshi families were approached, four consented and three completed GRACE; one withdrew before intervention delivery following hospital admission of the person living with dementia. Two families completed six sessions and one completed five; all received a complete intervention as defined in the core components. Delivery varied between families in language, use of interpretation and mode of delivery (Table 3).

**Table 3.** Variation in GRACE pilot delivery, personalised goals and learning informing further adaptation.

|  | **Family 1** | **Family 2** | **Family 3** |
| --- | --- | --- | --- |
| **Participants in GRACE** | Wife caring for husband | Daughter caring for father | Wife caring for husband |
| **Language and mode of delivery** | Spoken Sylheti; interpreter-supported; face-to-face | English; remote; no interpreter | English with family carer; remote; no interpreter |
| **Sessions completed** | 6 | 5 | 6 |
| **Goals set** | 1. Improve sleep<br>2. Improve diet<br>3. Participate in social activity | 1. Participate in social activity<br>2. Carer able to have a break | 1. Reduce carer anxiety<br>2. Carer able to have a break<br>3. Participate in social activity |
| **Goal status at final session** | 1. Much better<br>2. No longer relevant<br>3. A bit better | 1. A bit better<br>2. Much better | 1. A bit better<br>2. A bit better<br>3. Much better |
| **Key learning for adaptation** | English-only materials difficult to use through interpreter; need for translated/multimodal materials and interpreter preparation | Demonstrated importance of individual language preference rather than assuming Bengali-language delivery | Linguistically accessible community opportunities affected ability to pursue goals |

Across the three families, eight personalised goals were identified (mean 2.7 per family), relating to sleep, diet, social participation and family-carer wellbeing. At final review, seven of the eight goals had improved from baseline: three were rated as ‘much better’ and four as ‘a bit better’; the remaining goal was no longer considered relevant (Table 3).

All three family carers who completed GRACE and the facilitator agreed or strongly agreed with all AIM items, indicating high acceptability.

Pilot delivery generated further learning that informed subsequent cultural and linguistic adaptation. In the family receiving interpreter support, the family carer described a positive experience of interpretation:

> *“There were no difficulties at all. All the interpreters were brilliant.”* [Pilot family carer 1]

However, the facilitator described difficulties using the English-language intervention materials during interpreter-supported sessions, with delivery relying predominantly on verbal explanation:

> *“We couldn’t really use the booklets… it would have been useful to have some sort of translated version… or some sort of digital video version.”* [Facilitator]

The facilitator also identified continuity and preparation of interpreters as potentially important for supporting GRACE’s collaborative, goal-directed approach:

> *“The interpreters are normally used to medical conversations… I think if the interpreter had more understanding or training around what GRACE was or what the ethos was… that could have helped.”* [Facilitator]

Experiences across the three families reinforced the heterogeneity in language and communication preferences identified through stakeholder engagement (Theme 2). Families received GRACE through different combinations of spoken Sylheti and English, face-to-face and remote delivery, and interpreter support (Table 3), reinforcing the importance of responding to individual preferences rather than assuming that all

Bangladeshi families would require the same approach.

Personalised goals also reflected culturally meaningful aspects of families’ everyday lives. One family, for example, identified reconnecting the person living with dementia with his local mosque and wider community as a social participation goal. Pilot delivery also highlighted how the availability of linguistically accessible community opportunities could affect families’ ability to pursue their goals:

> *“My husband cannot speak English, so it is hard to find activities or groups for him to join and socialise with other people… we don’t have anyone speaking Bengali in the community.”* [Pilot family carer 3]

These experiences indicated that although GRACE could support families to identify personally meaningful goals, opportunities to pursue some goals could remain constrained by the availability of culturally and linguistically accessible community resources.

#### Further cultural and linguistic refinement

Pilot delivery and focus-group findings reinforced that neither English-language materials nor a single written translation would address the range of language, literacy and communication preferences within Bangladeshi families. The co-adaptation group therefore agreed to develop standard written Bengali materials as an additional option alongside English-language resources. Translation also identified GRACE concepts for which direct translation did not convey the intended meaning, including setting goals, lasting power of attorney, and apathy. These were discussed with Bengali-speaking PPI contributors and adapted to prioritise meaning and cultural relevance rather than word-for-word equivalence.

Pilot learning also informed changes to the facilitator workforce. Bengali- and Sylheti-speaking facilitators were recruited to increase capacity for linguistically responsive recruitment and intervention delivery, responding to issues around both access and trust (Theme 1) and language and communication (Theme 2), while reducing reliance on interpreter-mediated sessions.

Written Bengali did not meet all identified communication needs, particularly where spoken Sylheti was preferred or literacy in written Bengali was limited. The group therefore agreed to develop spoken Sylheti versions of text-heavy intervention content as an additional format, accessible through QR codes embedded within the corresponding materials. These resources are publicly available on the <u>GRACE website</u>.

Pilot experience also led the group to modify facilitator training. A section on working across languages and cultures was added, including practical guidance for interpreter-mediated delivery. This included briefing interpreters before sessions about GRACE and session goals, clarifying facilitator and interpreter roles, supporting meaning-based rather than word-for-word interpretation, using short chunks of information, checking understanding and allowing additional time. Training also emphasised avoiding assumptions based on cultural background and using families’ own priorities and goals to guide intervention delivery.

#### Adaptations across the iterative process

Table 4 summarises adaptations identified across the iterative process using FRAME. Adaptations arose both before pilot delivery through core-component work and stakeholder engagement, and reactively through pilot experience and subsequent family-carer and PPI engagement.

**Table 4.** Adaptations to GRACE across the iterative adaptation process, documented using FRAME.

| **Adaptation** | **Description and rationale** | **Planned/**<br>**reactive** | **Fidelity** |
| --- | --- | --- | --- |
| **Core-components work/ pre-pilot** |  |  |  |
| **Goal setting integrated into the first session** | Collaborative goal setting was incorporated into the first GRACE session rather than undertaken separately, to streamline delivery in routine services while retaining personalised, goal-directed intervention. | Planned | Fidelity-consistent* |
| **Abbreviated goal setting procedure** | An abbreviated approach to GAS was adopted. Only the baseline (0) and expected (+1) outcome levels were initially defined, reducing complexity for facilitators while retaining structured goal setting and review. | Planned | Fidelity-uncertain* |
| **Staged identification of personalised goals** | Dyads and facilitators identified at least one personalised goal in the first session, with additional goals identified or refined in subsequent sessions where appropriate, reducing the burden of completing all goal setting at the outset. | Planned | Fidelity-uncertain* |
| **Flexible intervention dose** | The intervention was adapted to allow 4–8 sessions according to family need, rather than requiring a fixed number of sessions, to support flexible delivery in routine services. | Planned | Fidelity-uncertain* |
| **Shortened facilitator training** | The five-day training programme used in the original trial was shortened to three six-hour training days, recognising the existing skills and experience of staff delivering GRACE alongside routine service roles, compared with facilitators recruited specifically for the trial. | Planned | Fidelity-consistent* |

| Stakeholder focus groups / pre-pilot |  |  |  |
| --- | --- | --- | --- |
| **More culturally representative imagery** | Photographs and illustrations in participant-facing materials were revised to better represent Bangladeshi families and culturally relevant everyday activities. | Planned | Fidelity-consistent* |
| Pilot and post-pilot refinement |  |  |  |
| **Standard Bengali written materials** | Participant-facing materials were translated into standard written Bengali as an additional option alongside English-language materials, following pilot and family-carer feedback highlighting variation in language and communication preferences. | Reactive | Fidelity-consistent* |
| **Recruitment of Bengali- and Sylheti-speaking facilitators** | Bengali- and Sylheti-speaking facilitators were recruited following pilot learning to increase capacity for linguistically responsive recruitment and delivery and reduce reliance on interpreter-mediated sessions. | Reactive | Fidelity-consistent* |
| **Spoken Sylheti resources** | Spoken Sylheti versions of text-heavy sections of intervention materials were developed and made accessible using QR codes, providing an alternative where written English or standard Bengali materials were less accessible or preferred. | Reactive | Fidelity-consistent* |
| **Facilitator training on working across languages and cultures** | Facilitator training was expanded to address culturally responsive delivery and working effectively with interpreters, in response to learning from interpreter-supported pilot delivery. | Reactive | Fidelity-consistent* |

\* Fidelity-consistent indicates that the adaptation was not expected to alter the intervention's underlying functioning or effectiveness.
Fidelity-uncertain indicates that the effect of the adaptation on intervention functioning or effectiveness was unknown.

## Discussion

This study demonstrates how cultural and linguistic adaptations to a complex psychosocial dementia intervention were identified, developed and refined through a structured, co-adaptation and iterative process. Defining the core functions of GRACE-family provided a reference point for adaptation, while stakeholder engagement identified anticipated cultural, linguistic and contextual considerations and pilot delivery exposed practical challenges that were less apparent before implementation. Subsequent adaptations included standard Bengali written materials, spoken Sylheti resources, recruitment of Bengali- and Sylheti-speaking facilitators, and additional facilitator preparation for culturally responsive and interpreter-mediated delivery.

Previous psychosocial dementia interventions have been adapted for different cultural contexts, commonly addressing language, literacy, culturally relevant examples and methods of delivery (Akhtar et al., 2026; James et al., 2021; McDermott et al., 2025). Our findings are consistent with this broader conceptualisation of cultural adaptation (Bernal et al., 2009; James et al., 2021; Resnicow et al., 1999). Different sources of evidence informed adaptation at different stages: pre-pilot engagement identified anticipated considerations relating to language, literacy, stigma, trust and cultural relevance, while pilot delivery revealed additional practical challenges, particularly around using intervention materials during interpreter-mediated sessions. These findings informed subsequent changes to intervention materials, facilitator workforce and preparation, supporting conceptualisations of adaptation as a process in which intervention-context fit is iteratively refined as implementation generates new learning 29).

Stigma, trust and community networks shaped how participants described engagement with dementia support, consistent with previous research with South Asian communities (Carter et al., 2024; Herat-Gunaratne et al., 2020; Mukadam et al., 2015). These findings highlighted that cultural adaptation involved not only how GRACE was delivered, but how families were reached and engaged. Following pilot delivery, Bengali- and Sylheti-speaking facilitators were recruited to increase capacity for linguistically responsive recruitment and delivery. However, participants also emphasized considerable variation within the Bangladeshi community in language, literacy, family circumstances and preferred modes of support. Cultural adaptation should therefore avoid replacing one standardised model with another based on assumptions about shared cultural preferences.

Personalised goal setting may provide one mechanism through which an intervention can respond to this heterogeneity. Within the small pilot, collaborative goal setting could be implemented across different linguistic and delivery arrangements, and families identified personally and culturally meaningful priorities relating to social participation, attending the mosque and family-carer wellbeing. These findings suggest that the goal-directed structure of GRACE-family could accommodate culturally meaningful priorities without requiring culturally prescriptive goal-setting methods. This is consistent with the person-centred principles underpinning GRACE-family, whereby families define what outcomes are meaningful rather than these being predetermined by the intervention (Budgett et al., 2024; Cooper et al., 2024; Wyman et al., 2024).

Previous adaptation of another psychosocial intervention for South Asian family carers similarly highlighted the importance of attending to family relationships, cultural beliefs and language preferences (Rapaport et al., 2025).

Focus groups identified the importance of spoken Sylheti and variation in literacy and preferred written language, while pilot delivery demonstrated the limitations of relying on English-language materials during interpreter-mediated sessions. Subsequent refinement therefore included both standard written Bengali materials and spoken Sylheti resources. The translation process demonstrated that some concepts required meaning-based adaptation rather than direct word-for-word translation. Together, these findings illustrate the importance of considering spoken language, literacy and communication preferences at an individual and local-community level rather than using ethnicity as a proxy for a family’s preferred language or format.

These linguistic considerations are particularly relevant in East London, where many Bangladeshi families originate from the Sylhet region of north-eastern Bangladesh (Rajina, 2026). Sylheti is primarily used as a spoken language within this community, while standard Bengali is the established written language (Lawson & Sachdev, 2004). However, literacy and written-language preferences vary. Providing English and standard Bengali written materials alongside spoken Sylheti resources therefore offered different formats that could be selected according to individual language, literacy and communication preferences.

Pilot delivery also highlighted that access to an interpreter alone may be insufficient for delivering a collaborative psychosocial intervention. Much existing dementia literature has focused on the role of interpreters during diagnosis and cognitive assessment (Brijnath et al., 2026; Nielsen et al., 2024). Brijnath and colleagues developed a national dementia interpreter training programme in Australia, demonstrating the potential value of specialist training for interpreters and clinicians (Brijnath et al., 2026). Comparatively less attention has been given to interpreter-mediated delivery of longer-term psychosocial interventions. Our findings suggest that facilitator training for working with interpreters, and orientation of interpreters to the intervention’s purpose and approach, may be important components of culturally and linguistically responsive delivery.

The wider community context may also constrain the extent to which personalised interventions can address inequalities in dementia support. Although GRACE enabled families to identify personally meaningful goals, opportunities to pursue these depended partly on the availability of culturally and linguistically accessible community resources (Cheston et al., 2025; McDermott et al., 2025; Nair et al., 2026).

Although the original NIDUS-Family intervention was cost-effective in the randomised controlled trial (Isaaq et al., 2025), culturally and linguistically responsive delivery may introduce additional costs associated with translation and audiovisual resources, interpreter-supported delivery, and recruitment and training of linguistically diverse facilitators. These were not evaluated in the present study and warrant consideration in future implementation and evaluation.

A strength of this study was the combination of co-adaptation, stakeholder engagement and pilot implementation within a structured adaptation process. FRAME supported transparent documentation of adaptations made during implementation; these are often incompletely reported, limiting opportunities for replication and learning (Escoffery et al., 2019).

Several limitations should be acknowledged. All participants in the stakeholder focus groups were women, as were all family carers participating in the pilot. While the latter reflects the gendered nature of family caregiving, it limited the representation of male perspectives. The family-carer focus group was small, with five of six participants being daughters-in-law, and recruitment through the community network of one Bengali PPI contributor may have limited the diversity of perspectives represented. The pilot involved only four families, three of whom completed the intervention, and was designed to inform intervention refinement rather than evaluate effectiveness; improvement across seven of eight personalised goals should therefore be interpreted descriptively. The study was conducted within a single East London memory service serving a large Sylheti-speaking Bangladeshi community, and further adaptation may be required in other geographical and service contexts. People living with dementia were also less well represented in the qualitative feedback than family carers. Finally, further evaluation is needed to understand the effects of these adaptations on intervention delivery, engagement, fidelity and outcomes.

## Conclusion

This study demonstrates the value of a structured, co-produced and iterative approach to culturally and linguistically adapting a complex psychosocial dementia intervention. Combining stakeholder engagement with pilot delivery enabled anticipated adaptation needs to be supplemented by practical learning that emerged through implementation. Adaptations addressed intervention materials, language and communication formats, facilitator workforce and training, while variation within the Bangladeshi community highlighted the importance of responding to individual language, literacy and cultural preferences rather than assuming homogeneous needs. This approach may provide a useful exemplar for adapting other complex psychosocial interventions where responsiveness to heterogeneous cultural and linguistic needs must be balanced with retention of core intervention functions.

## Data Availability

All data produced in the present study are available upon reasonable request to the authors

## Acknowledgments

We thank the INSTILLED study participants and the INSTILLED public and patient involvement group, whose generous time and commitment make our studies possible.

## References

Aarons, G. A., Hurlburt, M., & Horwitz, S. M. (2011). Advancing a Conceptual Model of Evidence-Based Practice Implementation in Public Service Sectors. Administration and Policy in Mental Health and Mental Health Services Research, 38(1), 4–23. 10.1007/s10488-010-0327-7

Akhtar, A., Nwofe, E., Dodd, E., Fry, G., Knight, N., Jutla, K., Cheston, R., & Parveen, S. (2026). Culturally adapted post-diagnostic dementia support for South Asian people living with dementia and caregivers: A rapid review. Age and Ageing, 55(9), afag266. 10.1093/ageing/afag266

Armstrong, M. J., Rastgardani, T., Gagliardi, A. R., & Marras, C. (2019). Barriers and facilitators of communication about off periods in Parkinson’s disease: Qualitative analysis of patient, carepartner, and physician Interviews. PLoS One, 14(4), e0215384. (30998707). 10.1371/journal.pone.0215384

Bernal, G., Jiménez-Chafey, M. I., & Domenech Rodríguez, M. M. (2009). Cultural adaptation of treatments: A resource for considering culture in evidence-based practice. Professional Psychology: Research and Practice, 40(4), 361–368. 10.1037/a0016401

Bhanu, C., Jones, M. E., Walters, K., Petersen, I., Manthorpe, J., Raine, R., Mukadam, N., & Cooper, C. (2020). Physical health monitoring in dementia and associations with ethnicity: A descriptive study using electronic health records. BJGP Open, 4(4), bjgpopen20X101080. 10.3399/bjgpopen20X101080

Braun, V., & Clarke, V. (2019). Reflecting on reflexive thematic analysis. *Qualitative Research in Sport*, Exercise and Health, 11(4), 589–597. 10.1080/2159676X.2019.1628806

Brijnath, B., Clarke, N., Markusevska, S., Garcia, E. G., Hlavac, J., Low, L.-F., Woodward-Kron, R., Antoniades, J., Enticott, J., Gilbert, A. S., LoGiudice, D., Lin, X., Hwang, K., White, J., & Cavuoto, M. (2026). Implementing interpreter training for dementia assessments at national scale using the RE-AIM framework: A mixed-methods evaluation. BMC Geriatrics, *26*(1), 516. 10.1186/s12877-026-07238-8

Budgett, J., Sommerlad, A., Kupeli, N., Zabihi, S., Rockwood, K., & Cooper, C. (2024). Personalized goals of people living with dementia and family carers: A content analysis of goals set within an individually tailored psychosocial intervention trial. Alzheimer’s & Dementia, 10(3), e12493. 10.1002/trc2.12493

Carnell Farrar. (2024). The economic impact of dementia. Alzheimer’s Society. https://www.alzheimers.org.uk/sites/default/files/2024-05/the-annual-costs-of-dementia.pdf

Carter, C., Roche, M., Whitfield, E., Budgett, J., Morgan-Trimmer, S., Zabihi, S., Birks, Y., Walter, F., Wilberforce, M., Jiang, J., Ahmed, R., Dowridge, W., Marshall, C. R., & Cooper, C. (2024). Equality of opportunity for timely dementia diagnosis (EQUATED): A qualitative study of how people from minoritised ethnic groups experience the early symptoms of dementia and seek help. Age and Ageing, 53(11), afae244. 10.1093/ageing/afae244

Cheston, R., Dodd, E., Smith, P., Woodstoke, N. S., Jutlla, K., Fry, G., Truswell, D., Butt, J., & Parveen, S. (2025). ‘You just can’t do that in dementia care’: Barriers to partnership working within dementia services for people from south Asian communities. Dementia, 24(4), 611–630. 10.1177/14713012241283189

Cooper, C., Vickerstaff, V., Barber, J., Phillips, R., Ogden, M., Walters, K., Lang, I., Rapaport, P., Orgeta, V., Rockwood, K., Banks, S., Palomo, M., Butler, L. T., Lord, K., Livingston, G., Banerjee, S., Manthorpe, J., Dow, B., Hoe, J., … Budgett, J. (2024). A psychosocial goal-setting and manualised support intervention for independence in dementia (NIDUS-Family) versus goal setting and routine care: A single-masked, phase 3, superiority, randomised controlled trial. The Lancet. Healthy Longevity, 5(2), e141–e151. 10.1016/S2666-7568(23)00262-3

de Boer, J., Longworth, G. R., Delfmann, L. R., Belmon, L. S., Vogelsang, M., Erikowa-Orighoye, O., An, Q., Deforche, B., Cardon, G., Verloigne, M., Altenburg, T., & Giné-Garriga, M. (2025). Exploring co-adaptation for public health interventions: Insights from a rapid review and interviews. BMC Public Health, 25(1), 614. 10.1186/s12889-025-21544-7

Department of Health and Social Care. (2025). Fit for the future: 10 Year Health Plan for England (CP 1350). https://www.gov.uk/government/publications/10-year-health-plan-for-england-fit-for-the-future

Escoffery, C., Lebow-Skelley, E., Udelson, H., Böing, E. A., Wood, R., Fernandez, M. E., & Mullen, P. D. (2019). A scoping study of frameworks for adapting public health evidence-based interventions. Translational Behavioral Medicine, 9(1), 1–10. 10.1093/tbm/ibx067

Herat-Gunaratne, R., Cooper, C., Mukadam, N., Rapaport, P., Leverton, M., Higgs, P., Samus, Q., & Burton, A. (2020). ‘In the Bengali Vocabulary, There Is No Such Word as Care Home’: Caring Experiences of UK Bangladeshi and Indian Family Carers of People Living With Dementia at Home. The Gerontologist, 60(2), 331–339. 10.1093/geront/gnz120

Isaaq, A., Cooper, C., Vickerstaff, V., Barber, J. A., Walters, K., Lang, I. A., Rapaport, P., Orgeta, V., Rockwood, K., Butler, L. T., Lord, K., Livingston, G., Banerjee, S., Manthorpe, J., Kales, H. C., Budgett, J., & Hunter, R. (2025). Cost-utility of a new psychosocial goal-setting and manualised support intervention for independence in dementia (NIDUS-Family) versus goal setting and routine care: An economic evaluation embedded within a randomised controlled trial. The Lancet Healthy Longevity, 6(2). 10.1016/j.lanhl.2024.100676

James, T., Mukadam, N., Sommerlad, A., Ceballos, S. G., & Livingston, G. (2021). Culturally tailored therapeutic interventions for people affected by dementia: A systematic review and new conceptual model. The Lancet Healthy Longevity, 2(3), e171–e179. 10.1016/S2666-7568(21)00001-5

Kishita, N., Hammond, L., Dietrich, C. M., & Mioshi, E. (2018). Which interventions work for dementia family carers?: An updated systematic review of randomized controlled trials of carer interventions. International Psychogeriatrics, 30(11), 1679–1696. 10.1017/S1041610218000947

Lawson, S., & Sachdev, I. (2004). Identity, Language Use, and Attitudes: Some Sylheti-Bangladeshi Data from London, UK. Journal of Language and Social Psychology, 23(1), 49–69. 10.1177/0261927X03261223

McDermott, O., Sobers, T., Mukadam, N., Lee, A. R., & Orrell, M. (2025). What Works Well for People With Dementia and Their Supporters From South Asian, African and Caribbean Communities in the UK: A Narrative Synthesis Systematic Review and Expert Consultations. International Journal of Geriatric Psychiatry, 40(3), e70047. 10.1002/gps.70047

Moore, G., Campbell, M., Copeland, L., Craig, P., Movsisyan, A., Hoddinott, P., Littlecott, H., O’Cathain, A., Pfadenhauer, L., Rehfuess, E., Segrott, J., Hawe, P., Kee, F., Couturiaux, D., Hallingberg, B., & Evans, R. (2021). Adapting interventions to new contexts-the ADAPT guidance. BMJ, 374, n1679. 10.1136/bmj.n1679

Mukadam, N., Marston, L., Lewis, G., & Livingston, G. (2022). Risk factors, ethnicity and dementia: A UK Biobank prospective cohort study of White, South Asian and Black participants. PloS One, 17(10), e0275309. 10.1371/journal.pone.0275309

Mukadam, N., Waugh, A., Cooper, C., & Livingston, G. (2015). What would encourage help-seeking for memory problems among UK-based South Asians? A qualitative study. 10.1136/bmjopen-2015-007990

Nair, P., Spencer, E., Walters, K., Stevenson, F., Dooley, J., & Davies, N. (2026). Key factors influencing post-diagnostic support and care planning for people with dementia from South Asian backgrounds: A systematic review of qualitative studies. BMC Geriatrics, 26, 205. 10.1186/s12877-026-07064-y NICE. (2018, June 20). Recommendations | Dementia: Assessment, management and support for people living with dementia and their carers | Guidance | NICE. https://www.nice.org.uk/guidance/ng97/chapter/Recommendations#managing-non-cognitive-symptoms

Nielsen, T. R., Franzen, S., Watermeyer, T., Jiang, J., Calia, C., Kjærgaard, D., Bothe, S., & Mukadam, N. (2024). Interpreter-mediated neuropsychological assessment: Clinical considerations and recommendations from the European Consortium on Cross-Cultural Neuropsychology (ECCroN). The Clinical Neuropsychologist, 38(8), 1775–1805. 10.1080/13854046.2024.2335113

Penfold, R. S., Wilkinson, T., Stirland, L. E., MacRae, C., Russ, T. C., Shenkin, S. D., Vardy, E., Anand, A., Guthrie, B., Sampson, E. L., & MacLullich, A. M. J. (2026). Prevalence and outcomes of recorded dementia vary by data source: A population cohort study of 133 407 older adults. Age and Ageing, 55(2), afag038. 10.1093/ageing/afag038

Prince, M., Knapp, M., Guerchet, M., McCrone, P., Prina, M., Comas-Herrera, A., Wittenberg, R., Adelaja, B., Hu, B., King, D., Rehill, A., & Salimkumar, D. (2014). Dementia UK: Second Edition - Overview. https://kclpure.kcl.ac.uk/portal/en/publications/dementia-uk-second-edition--overview(22d426a1-c0aa-4b3f-8fcb-1db4a3f4c4d7)/export.html

Rajina, F. (2026). From desh to bidesh: History of migration and activism of British Bangladeshi Muslims in East London. Identities, 0(0), 1–19. 10.1080/1070289X.2026.2641371

Rapaport, P., Burton, A., Leverton, M., Herat-Gunaratne, R., Beresford-Dent, J., Lord, K., Downs, M., Boex, S., Horsley, R., Giebel, C., & Cooper, C. (2020). “I just keep thinking that I don’t want to rely on people.” a qualitative study of how people living with dementia achieve and maintain independence at home: Stakeholder perspectives. BMC Geriatrics, 20(1), 5. 10.1186/s12877-019-1406-6

Rapaport, P., Muralidhar, M., Amador, S., Mukadam, N., Bhojwani, A., Beeson, C., & Livingston, G. (2025). Sleep Disturbances and Dementia in the UK South Asian Community: A Qualitative Study to Inform Future Adaptation of the DREAMS-START Intervention. Geriatrics, 10(5), 121. 10.3390/geriatrics10050121

Reardon, C. M., Damschroder, L. J., Ashcraft, L. E., Kerins, C., Bachrach, R. L., Nevedal, A. L., Domlyn, A. M., Dodge, J., Chinman, M., & Rogal, S. (2025). The Consolidated Framework for Implementation Research (CFIR) User Guide: A five-step guide for conducting implementation research using the framework. Implementation Science : IS, 20, 39. 10.1186/s13012-025-01450-7

Resnicow, K., Baranowski, T., Ahluwalia, J. S., & Braithwaite, R. L. (1999). Cultural sensitivity in public health: Defined and demystified. Ethnicity & Disease, 9(1), 10–21.

Tower Hamlets Partnership. (2023). Tower Hamlets: State of the borough 2023. https://www.towerhamlets.gov.uk/Documents/Borough_statistics/State-of-the-Borough.pdf

Turner-Stokes, L. (2009). Goal attainment scaling (GAS) in rehabilitation: A practical guide. Clinical Rehabilitation, 23(4), 362–370. 10.1177/0269215508101742

Weiner, B. J., Lewis, C. C., Stanick, C., Powell, B. J., Dorsey, C. N., Clary, A. S., Boynton, M. H., & Halko, H. (2017). Psychometric assessment of three newly developed implementation outcome measures. Implementation Science : IS, 12, 108. 10.1186/s13012-017-0635-3

Wiltsey Stirman, S., Baumann, A. A., & Miller, C. J. (2019). The FRAME: An expanded framework for reporting adaptations and modifications to evidence-based interventions. Implementation Science: IS, 14(1), 58. 10.1186/s13012-019-0898-y

Wyman, D., Butler, L. T., Morgan-Trimmer, S., Bright, P., Barber, J., Budgett, J., Walters, K., Lang, I., Rapaport, P., Banks, S., Palomo, M., Orgeta, V., Livingston, G., Rockwood, K., Lord, K., Manthorpe, J., Dow, B., Hoe, J., & Cooper, C. (2024). Process evaluation of a New psychosocial goal-setting and manualised support intervention for Independence in Dementia (NIDUS-Family). Age and Ageing, 53(8), afae181. 10.1093/ageing/afae181

